# Stunting, Thinness, Obesity, and Double Burden of Malnutrition in Vietnamese Urban Children

**DOI:** 10.64898/2026.07.26.26358982

**Authors:** Nhan Thi Ho, Michelle Hermiston, Minh Anh Nguyen, Quyet Van Nguyen, Anh Quoc Dao, Chi Thi Linh Tran, Huong Thi Minh Le, Thuy Van Pham, Lan Tuyet Phung, Hoan Thi Nguyen, Quang Ngoc Nguyen, Lam Nam Phung, Cuong Tat Do, An Nhat Pham

**Affiliations:** Research Management Department, Vinmec International Hospital, Hanoi, Vietnam; College of Health Sciences, VinUniversity, Hanoi, Vietnam; Health Check Department, Vinmec Times City International Hospital, Vinmec International Hospital, Hanoi, Vietnam; Pediatric Department, Vinmec Central Park International Hospital, Vinmec International Hospital, Hanoi, Vietnam; Pediatric Department, Vinmec Haiphong International Hospital, Vinmec International Hospital, Haiphong, Vietnam; Pediatric Department, Vinmec Times City International Hospital, Vinmec International Hospital, Hanoi, Vietnam; Nutrition Department, Vinmec Times City International Hospital, Vinmec International Hospital, Hanoi, Vietnam; Vinmec Riverside International Hospital, Vinmec International Hospital, Hanoi, Vietnam

**Keywords:** stunting, thinness, obesity, double burden of malnutrition, secular trends, Vietnam, school-age children, nutrition transition, generalized estimating equations

## Abstract

**Objectives:** To characterize prevalence and secular trends in stunting, thinness, overweight, obesity, and the double burden of malnutrition (DBM) in urban Vietnamese children.

**Methods:** We analyzed 249,745 annual health check visits from 97,111 children aged 18 months to 18 years attending a private school system in Hanoi, Ho Chi Minh City, and Haiphong from 2018 to 2025. Temporal trends in stunting, thinness, and obesity were modelled using generalized estimating equation (GEE) logistic regression.

**Results:** A total of 51,034 males and 46,077 females were included. Stunting prevalence was low but rose significantly in both sexes (males: 0.7% to 1.3%, p=0.006, females: 0.7% to 1.7%, p<0.001), concentrated in children under 10 years (GEE OR/year=1.076 and 1.089, both p<0.05), females exceeded males by 2025. Obesity declined overall in males (21.4% to 17.0%) and females (6.2% to 5.0%, both p<0.001), but rose significantly among males aged 15-18 years (OR=1.055, p=0.021). Overweight increased in males (19.7% to 20.5%, p=0.010) but declined in females (18.0% to 14.8%, p<0.001). Individual-level DBM (concurrent stunting with overweight/obesity) was uncommon (<0.2%). City-level heterogeneity was marked, with Haiphong showing the highest male obesity (22.7% in 2025) and individual-level DBM.

**Conclusion:** Vietnamese urban schoolchildren experienced simultaneous, divergent nutritional burdens from 2018-2025: rising stunting and thinness under age 10, declining early childhood obesity, and rising adolescent male obesity. Individual-level DBM was rare, but a population-level double burden was evident across age groups. City, age, and sex specific nutrition and school health policies are needed to address this complex malnutrition transition.

## INTRODUCTION

Child malnutrition in its multiple forms remains one of the foremost preventable threats to human health, development, and economic productivity worldwide. Globally, an estimated 148 million children under five years of age were stunted in 2022, while approximately 45 million suffered from wasting, representing both a humanitarian burden and a major constraint on human capital formation across low- and middle-income countries (LMICs) (1,2). A pooled analysis of nationally representative surveys from 62 LMICs documented a stunting prevalence of 29.1%, wasting of 6.3%, and underweight of 13.7% among children, with Southern and Southeastern Asia accounting for a disproportionately large share of the global burden (1). Despite decades of programmatic investment, progress has been uneven: while stunting has declined in many middle-income countries, absolute numbers of affected children remain high, and reductions have been slowest in the most marginalized populations, including ethnic minorities, rural households, and children from the poorest wealth quintiles (2,3).

Simultaneously across Asia, a rapid nutrition transition is superimposing overweight and obesity onto populations where undernutrition has not yet been resolved, producing the phenomenon known as the *double burden of malnutrition* (DBM). The DBM manifests at the population level as the coexistence of undernutrition and overnutrition across age groups within the same community, and at the individual level as concurrent stunting and overweight or obesity within the same child (4,5). In Southeast Asia, projections to 2030 indicate that while stunting and thinness among children aged 5 to 19 years are expected to decline (from 37% to 27% and from 30% to 6%, respectively), overweight prevalence is projected to nearly triple (from 6% to 17%) and obesity to more than double (from 3% to 10%) (6), signaling that the region faces not a resolution of malnutrition but a transformation of its form. This triple burden which encompasses undernutrition, overnutrition, and micronutrient deficiency has been systematically documented across Indonesia, Malaysia, Thailand, and Vietnam, with studies reporting concurrent anemia in 25%, iron deficiency in 14%, vitamin A deficiency in 6%, and vitamin D deficiency in 40% of children in affected countries (7).

The individual-level DBM, defined as concurrent stunting and overweight or obesity in the same child, represents a particularly challenging clinical and public health phenomenon, as the two conditions share limited etiological overlap and may require contradictory intervention approaches (4,5). Data from the Young Lives longitudinal cohort which followed children in India, Peru, and Vietnam demonstrated that childhood and adolescence are critical windows during which transitions between normal nutritional status, stunting, overweight, and concurrent stunting and overweight (CSO) occur, and that reversal from the combined CSO state to normal nutrition is rare once established (8). A systematic review of coexisting forms of malnutrition (CFM) in children under five across South and Southeast Asia found CFM prevalence reached 23% in some settings, with higher maternal education and male sex associated with lower odds (11).

Vietnam, over the past two decades, has achieved remarkable reductions in child stunting: national under-five stunting declined from 41.5% in 2000 to 19.3% by 2020, driven by rising household wealth, improved maternal education, expanded sanitation coverage, and targeted multisectoral nutrition programs encompassing micronutrient supplementation, school-based milk provision, community health workers, and poverty reduction strategies (12). Analyses from the Young Lives cohort confirmed that Vietnamese children had the lowest stunting burden among four LMIC cohort countries (Ethiopia, India, Peru, Vietnam), with parental education and household wealth index identified as the strongest protective determinants (13). Nevertheless, stunting and thinness remain concentrated among ethnic minority groups, children in mountainous and peri-urban areas, and the lowest socioeconomic quintiles, while overweight and obesity have surged in urban schoolchildren, particularly in Hanoi and Ho Chi Minh City where prevalence now exceeds 30 to 40% (14,15).

Despite this growing body of evidence, several critical gaps persist. First, the vast majority of published studies characterizing stunting, thinness, and DBM trends in Vietnam and Southeast Asia have used cross-sectional or single time-point data, limiting the ability to characterize true secular trends (15,16). Second, very few studies have simultaneously examined all three dimensions of child nutritional status (stunting, thinness, and the double burden) within the same cohort, across the full pediatric age range from infancy through late adolescence, and across multiple cities (7,11). Third, city-level heterogeneity within Vietnam has rarely been assessed with adequate statistical power and comparative longitudinal data across cities are lacking.

This study analyzed the annual school health check data of children age from 18 months to 18 years attending a private school system in 3 major cities Hanoi, Hochiminh and Haiphong in Vietnam. The aims are to: (1) estimate the annual prevalence of stunting, thinness, overweight, obesity, and the double burden of malnutrition from 2018 to 2025, stratified by sex, age group, and city; (2) examine secular trends in each nutritional outcome adjusting for age group, city, and a calendar year×age interaction; and (3) characterize temporal trends in the individual-level double burden (concurrent stunting with overweight or obesity). This study aims to provide an evidence base for targeting child growth interventions and school health policies in Vietnam and comparable LMICs.

## METHODS

### Study design and setting

This study analyzed the annual school health check data of 249,745 child-year visits of 97,111 unique children aged from 18 months to 18 years attending a private school system in 3 major cities Hanoi, Hochiminh and Haiphong in Vietnam. The present analysis uses examination records collected between 2018 and 2025 inclusive. The 2021 data was not available due to COVID-19 restriction.

This research was conducted in accordance with the Declaration of Helsinki. Ethical approval was granted by Vinmec Ethical Committee (approval number 0231/2024/CN/HDDD VMEC). As the study used de-identified routinely collected health data from school health examinations, a waiver of individual informed consent was approved by the ethics committee.

### Participants

All children aged 18 months to 18 years with at least one recorded height and weight measurement were included. Observations were excluded if age, height, or weight was missing or biologically implausible. Each child-year combination constituted an analytical observation for the repeated cross-sectional prevalence analyses. Children could contribute multiple observations across calendar years.

### Anthropometric classification against who reference standards

Anthropometric indices were classified according to WHO reference standards, applied separately by age group and sex. For children aged under 5 years, height-for-age (HFA) was evaluated against the WHO Child Growth Standards (2006) and adiposity was evaluated against weight-for-height (WFH) z-score reference tables (17). For children aged 5 to 18 years, HFA and BMI-for-age were evaluated against the WHO Reference 2007 (18).

For children aged under 5 years, HFA status was classified as: stunting (HFA < −2 SD), normal HFA (HFA >= −2 SD and <= +3 SD), extreme tallness (HFA > +3 SD). WFH status for under-5 years was classified as: thinness (WFH < −2 SD), normal (WFH >= −2 SD and <= +2 SD), overweight (WFH > +2 SD and <= +3 SD), obesity (WFH > +3 SD).

For children aged 5 to 18 years, HFA were classified using the same SD thresholds. BMI-for-age was classified as: thinness (BMI-for-age < −2 SD), normal (BMI-for-age >= −2 SD and <= +1 SD), overweight (BMI-for-age > +1 SD and <= +2 SD), obesity (BMI-for-age > +2 SD).

### Definition of double burden of malnutrition

The double burden of malnutrition (DBM) was defined at the individual level by simultaneously classifying each child’s HFA status and BMI/WFH status, with focus on these categories: 1. Stunting + Obesity; 2. Stunting + Overweight; 3. Stunting + Thinness. For primary DBM analyses, children exhibiting the category of greatest clinical and public-health concern which is stunting coexisting with overweight or obesity (Stunting + OW/OB) were additionally analyzed as a binary indicator. The prevalence of each DBM category was computed as the proportion of all observations with non-missing data for both HFA and BMI/WFH classification in a given calendar year, stratified by sex.

### Statistical analysis

All statistical analyses were conducted in R version 4.5.1 (19). A two-tailed alpha level of 0.05 was used as the threshold for statistical significance.

### Prevalence Estimation

The prevalence of stunting, thinness, overweight, obesity, and each DBM category was estimated as a proportion with simultaneous 95% confidence intervals (CIs) using the Sison-Glaz method for multinomial proportions as implemented in the DescTools package (20). Prevalence estimates and their 95% CIs were computed separately by calendar year, sex, age group, and city.

### Temporal Trend Analysis

Simple temporal trends in annual prevalence over the 2018-2025 study period were tested using the Cochran-Armitage test for trend in proportions. To model population-average secular trends of stunting, thinness, and obesity while accounting for the repeated observations of individual children across years, we fitted generalized estimating equation (GEE) models using the geepack package (version 1.3.13) (21). GEE models were specified with a logit link function (binomial family) and an independence working correlation structure.

All GEE models included calendar year (modelled as a linear integer covariate, centered at 2018), age group (four categories: <5 year, 5 to <10 year, 10 to <15 year, 15 to 18 year), city as fixed covariates. An interaction term between calendar year and age group was included to test whether secular trends in nutritional status differed by developmental stage. Separate models were fitted for males and females. Results are presented as odds ratios (ORs) per additional calendar year with 95% CIs, derived from estimated marginal slopes using the emmeans package (22).

## RESULTS

### Study Population

A total of 249,745 child-year visits of 97,111 unique children from 2018 to 2025 were included in the analysis, comprising 129,955 visits in 51,034 males and 119,790 visits in 46,077 females. Sample sizes increased substantially from 2018 (males: 3,546, females: 3,368) to 2025 (males: 24,670, females: 22,943). Data were drawn from three cities: Hanoi (HHN), which contributed the largest share of observations across all years, Ho Chi Minh City (HCP), and Haiphong (HHP). The full year-by-year, city-stratified sample sizes are presented in **Supplementary Materials**.

### Secular Trends in Stunting

Stunting prevalence was low overall but showed statistically significant increasing trends across the study period in both sexes (Cochran-Armitage test: males *p* = 0.006, females *p* < 0.0001) (**Table 1**, **Table 2, Supplementary Materials**). In males, stunting rose from 0.7% (95%CI = 0.4, 1.1%) in 2018 to 1.3% (1.2, 1.4%) in 2025, while in females it rose from 0.7% (0.4, 1.0%) to 1.7% (1.5, 1.8%) over the same period. By 2025, females had a consistently higher stunting prevalence than males, a reversal of the near-equal prevalence seen in 2018 (**Figure 1**, **Table 1**, **Table 2**). Age-stratified analyses (**Figure 2**) revealed that the rising stunting trend was most pronounced in the under 5 and 5 to <10-year age groups for both sexes, while the 15 to 18-year group showed negligible change. The GEE model confirmed these age-group differences: among males, the OR for stunting per additional calendar year was 1.076 (95%CI = 1.039, 1.116) in the under 5 age group and 1.053 (1.002, 1.108) in the 5 to <10 year group, but was non-significant in older age groups. In females, the corresponding ORs were 1.089 (95%CI = 1.047, 1.134) and 1.086 (95%CI = 1.025, 1.150) for the same age groups (**Table 2**, **Figure 1**).

**Figure 1.**
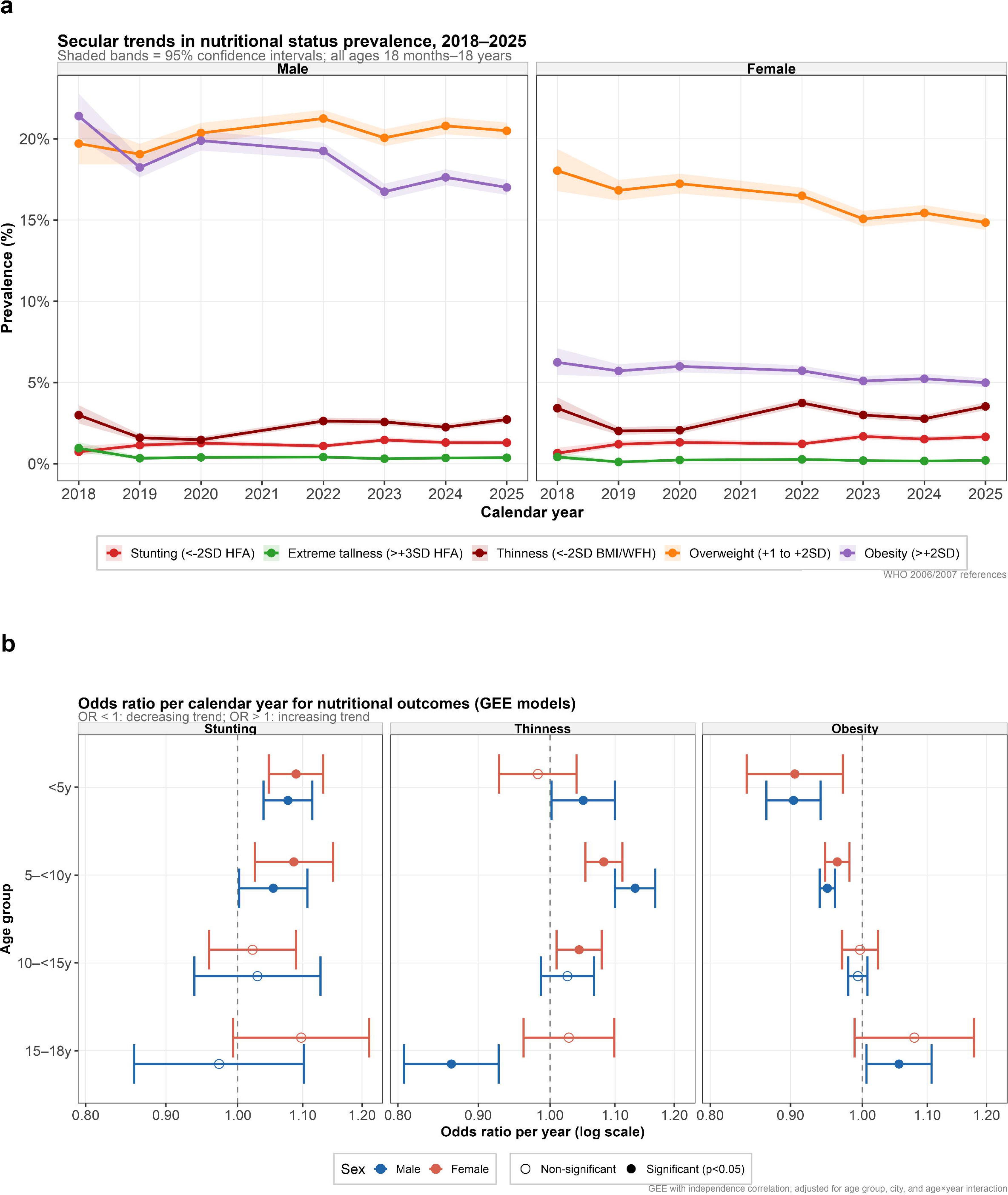
(a). Secular trends in the prevalence of stunting (HFA < −2 SD), thinness (BMI/WFH <-2 SD), overweight (+1 to +2 SD), and obesity (> +2 SD) from 2018 to 2025, stratified by sex (males left, females right). Lines represent annual point prevalence estimates, shaded bands represent simultaneous 95% confidence intervals (Sison-Glaz method). All ages 18 months to 18 years. (b). Forest plot of GEE-derived odds ratios per additional calendar year. For stunting (left), thinness (center), and obesity (right) by age group and sex. Filled circles = statistically significant (p < 0.05), open circles = non-significant. OR < 1 indicates a decreasing trend, OR > 1 indicates an increasing trend. Horizontal bars = 95% CIs. Models adjusted for city and year×age-group interaction.

**Figure 2.**
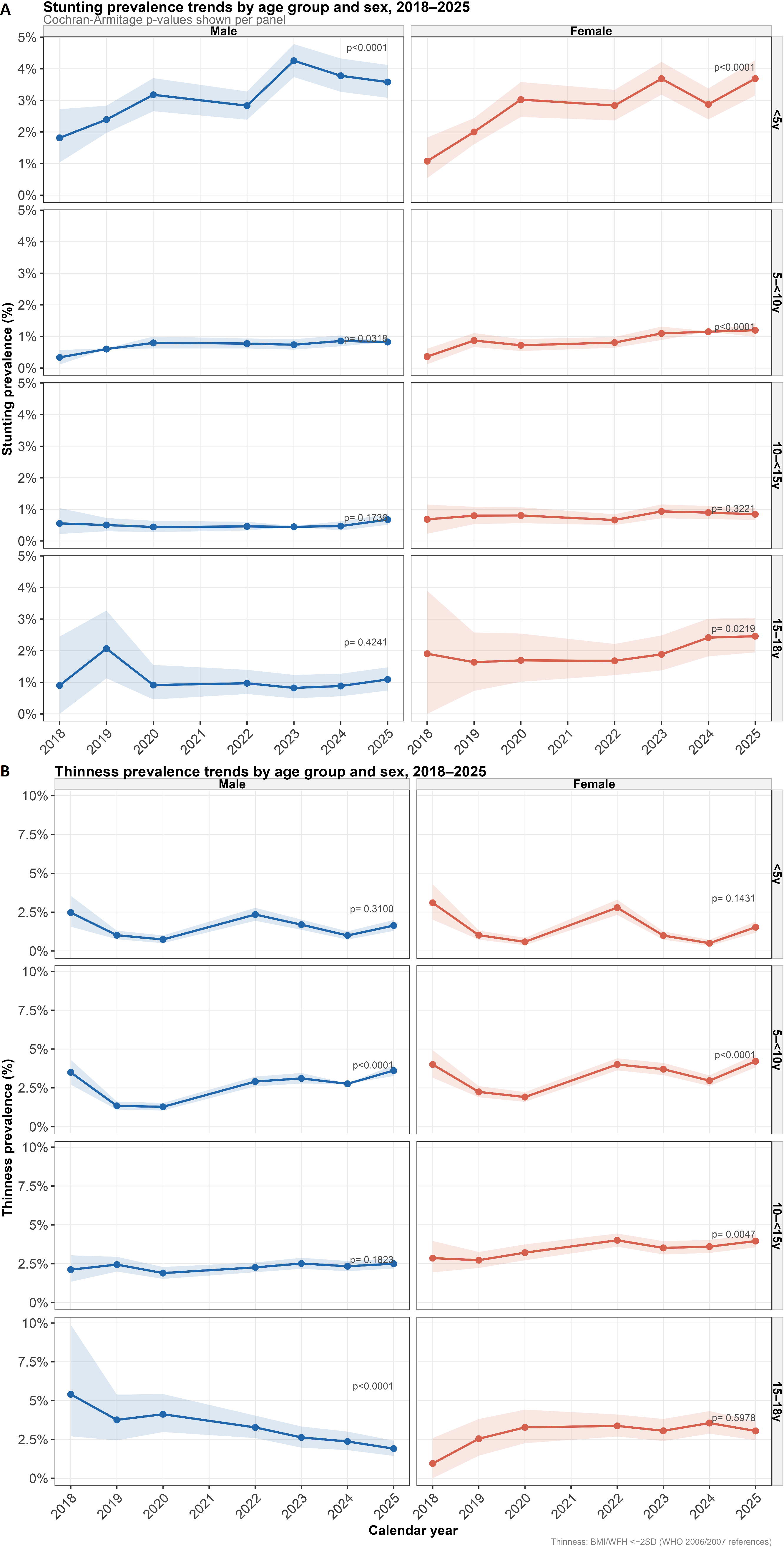
Stunting and thinness prevalence trends by age group (rows) and sex (columns), 2018 to 2025. Cochran-Armitage trend test p-values are annotated per panel. Lines and shaded bands represent prevalence estimates and 95% CIs respectively. A: Stunting, B: Thinness.

**Table 1.**
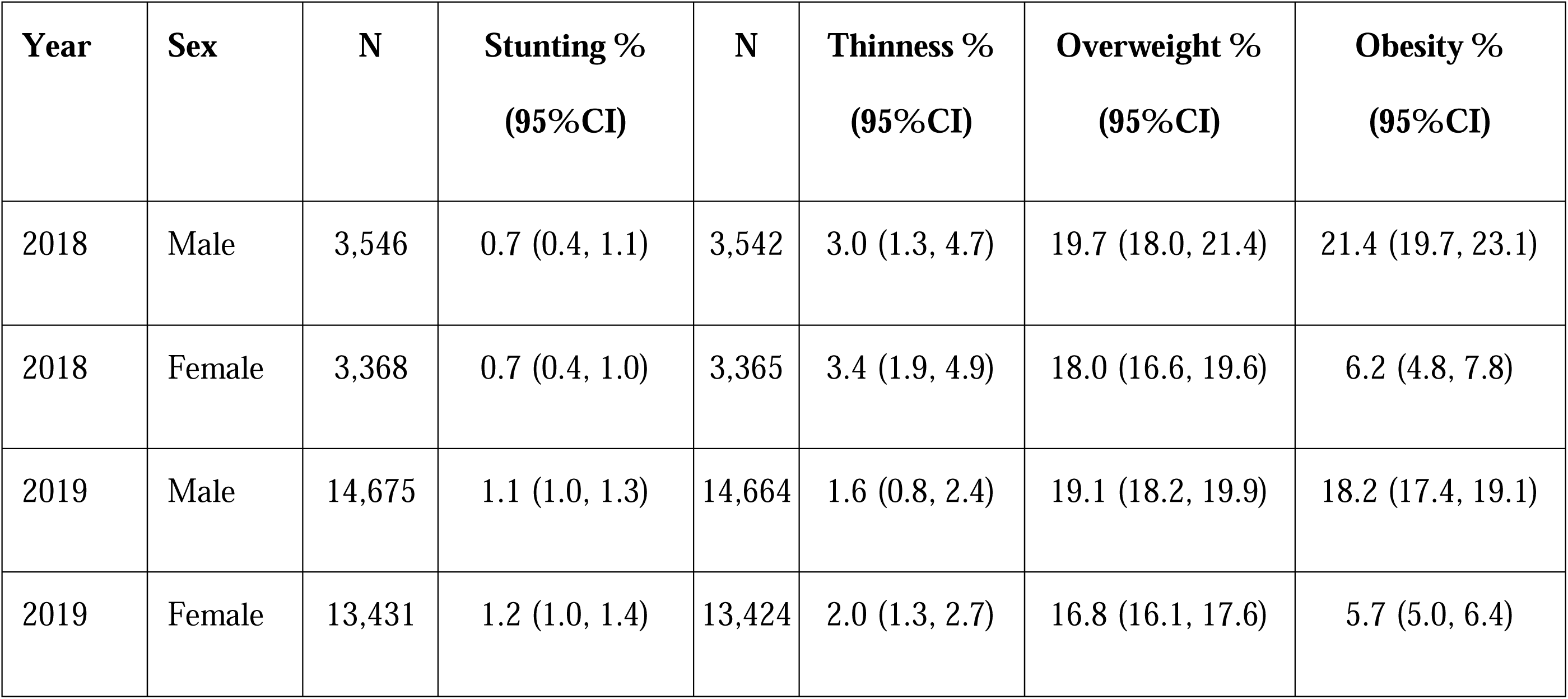
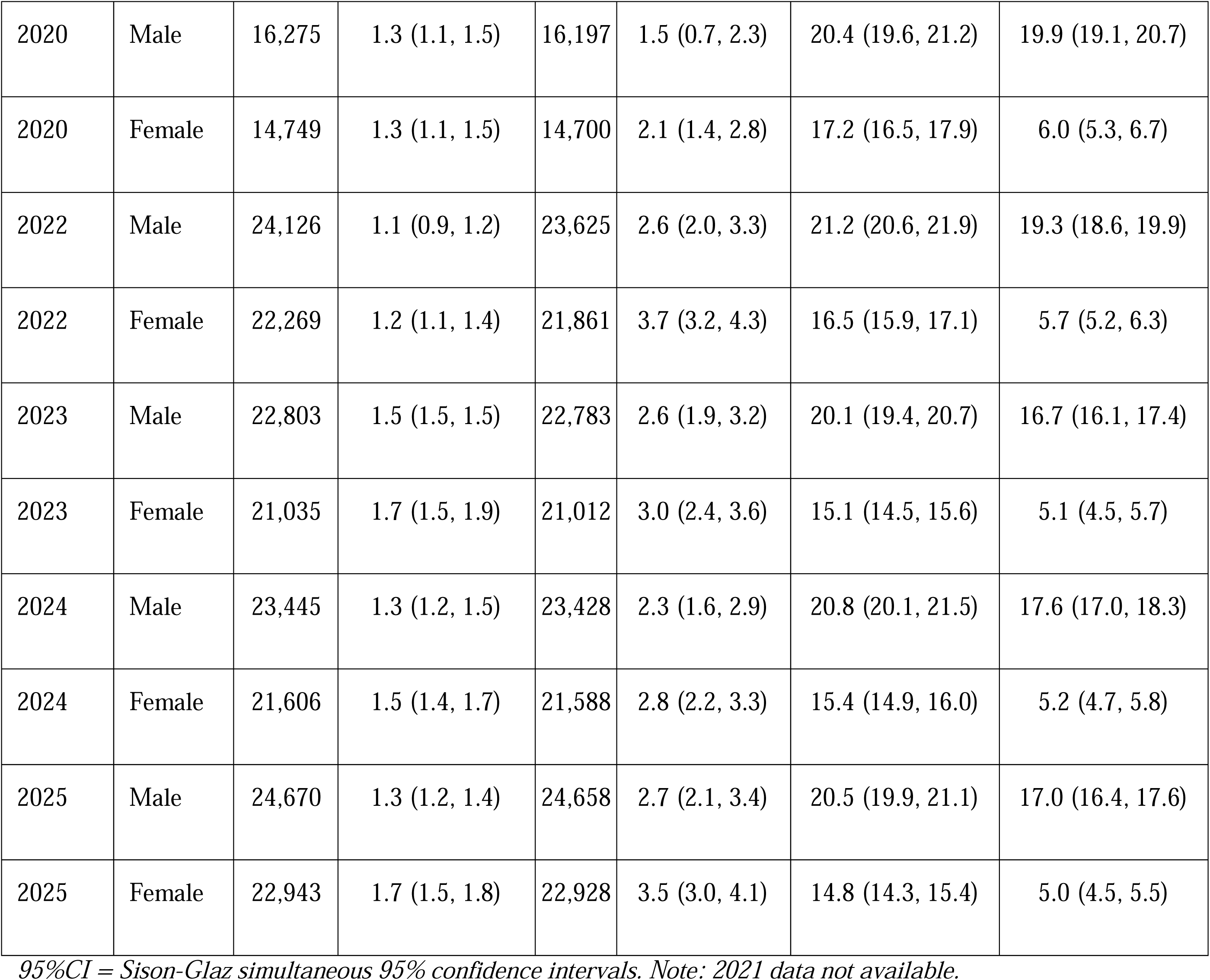
Stunting, thinness, overweight, obesity prevalence (%, 95%CI) by calendar year and sex, from 2018 to 2025.

| Year | Sex | N | Stunting %<br>(95%CI) | N | Thinness %<br>(95%CI) | Overweight %<br>(95%CI) | Obesity %<br>(95%CI) |
| --- | --- | --- | --- | --- | --- | --- | --- |
| 2018 | Male | 3,546 | 0.7 (0.4, 1.1) | 3,542 | 3.0 (1.3, 4.7) | 19.7 (18.0, 21.4) | 21.4 (19.7, 23.1) |
| 2018 | Female | 3,368 | 0.7 (0.4, 1.0) | 3,365 | 3.4 (1.9, 4.9) | 18.0 (16.6, 19.6) | 6.2 (4.8, 7.8) |
| 2019 | Male | 14,675 | 1.1 (1.0, 1.3) | 14,664 | 1.6 (0.8, 2.4) | 19.1 (18.2, 19.9) | 18.2 (17.4, 19.1) |
| 2019 | Female | 13,431 | 1.2 (1.0, 1.4) | 13,424 | 2.0 (1.3, 2.7) | 16.8 (16.1, 17.6) | 5.7 (5.0, 6.4) |
| 2020 | Male | 16,275 | 1.3 (1.1, 1.5) | 16,197 | 1.5 (0.7, 2.3) | 20.4 (19.6, 21.2) | 19.9 (19.1, 20.7) |
| 2020 | Female | 14,749 | 1.3 (1.1, 1.5) | 14,700 | 2.1 (1.4, 2.8) | 17.2 (16.5, 17.9) | 6.0 (5.3, 6.7) |
| 2022 | Male | 24,126 | 1.1 (0.9, 1.2) | 23,625 | 2.6 (2.0, 3.3) | 21.2 (20.6, 21.9) | 19.3 (18.6, 19.9) |
| 2022 | Female | 22,269 | 1.2 (1.1, 1.4) | 21,861 | 3.7 (3.2, 4.3) | 16.5 (15.9, 17.1) | 5.7 (5.2, 6.3) |
| 2023 | Male | 22,803 | 1.5 (1.5, 1.5) | 22,783 | 2.6 (1.9, 3.2) | 20.1 (19.4, 20.7) | 16.7 (16.1, 17.4) |
| 2023 | Female | 21,035 | 1.7 (1.5, 1.9) | 21,012 | 3.0 (2.4, 3.6) | 15.1 (14.5, 15.6) | 5.1 (4.5, 5.7) |
| 2024 | Male | 23,445 | 1.3 (1.2, 1.5) | 23,428 | 2.3 (1.6, 2.9) | 20.8 (20.1, 21.5) | 17.6 (17.0, 18.3) |
| 2024 | Female | 21,606 | 1.5 (1.4, 1.7) | 21,588 | 2.8 (2.2, 3.3) | 15.4 (14.9, 16.0) | 5.2 (4.7, 5.8) |
| 2025 | Male | 24,670 | 1.3 (1.2, 1.4) | 24,658 | 2.7 (2.1, 3.4) | 20.5 (19.9, 21.1) | 17.0 (16.4, 17.6) |
| 2025 | Female | 22,943 | 1.7 (1.5, 1.8) | 22,928 | 3.5 (3.0, 4.1) | 14.8 (14.3, 15.4) | 5.0 (4.5, 5.5) |

95%CI = Sison-Glaz simultaneous 95% confidence intervals. Note: 2021 data not available.

**Table 2.** Secular trends of stunting, thinness, and obesity by age group and sex.

| Outcome | Age group | Males OR/yr (95% CI)\$ | Females OR/yr (95% CI)\$ |
| --- | --- | --- | --- |
| Stunting | <5 years | 1.076 (1.039, 1.116)* | 1.089 (1.047, 1.134)* |
|  | 5 to <10 years | 1.053 (1.002, 1.108)* | 1.086 (1.025, 1.150)* |
|  | 10 to <15 | 1.029 (0.938, 1.129) | 1.022 (0.959, 1.089) |
|  | years |  |  |
|  | 15 to 18 years | 0.973 (0.859, 1.102) | 1.098 (0.993, 1.213) |
| Thinness | <5 years | 1.050 (1.002, 1.100)* | 0.982 (0.928, 1.040) |
|  | 5 to <10 years | 1.133 (1.100, 1.167)* | 1.082 (1.053, 1.112)* |
|  | 10 to <15 years | 1.026 (0.987, 1.067) | 1.044 (1.010, 1.079)* |
|  | 15 to 18 years | 0.865 (0.807, 0.928)* | 1.028 (0.962, 1.099) |
| Obesity | <5 years | 0.904 (0.869, 0.941)* | 0.906 (0.844, 0.972)* |
|  | 5 to <10 years | 0.950 (0.939, 0.961)* | 0.964 (0.947, 0.981)* |
|  | 10 to <15 years | 0.994 (0.980, 1.008) | 0.997 (0.971, 1.023) |
|  | 15 to 18 years | 1.055 (1.006, 1.107)* | 1.079 (0.989, 1.178) |

*\$OR/yr = odds ratio per additional calendar year; \* = $p < 0.05$ . GEE model with logit link, independence correlation structure, adjusted for age group, city, and year × age group interaction; fitted separately by sex. $OR < 1$ = decreasing annual trend; $OR > 1$ = increasing trend. # Cochran–Armitage test for linear trend in proportions stratified by city.*

City-specific trend tests (**Supplementary Materials**) showed that the rising stunting trend was statistically significant in Hanoi (*p* < 0.0001) and Ho Chi Minh City (*p* = 0.019), but not in Haiphong (*p* = 0.961). The stacked city figures (**Figures 3**) illustrate that Haiphong and Hanoi consistently had the higher stunting prevalence throughout the study period (reaching 1.5% in males and 1.8% in females in Haiphong and 1.6% and 1.9% in Hanoi by 2025, respectively) as compared to Ho Chi Minh City (0.6% and 1.0% in, respectively) (**Supplementary Material**).

**Figure 3.**
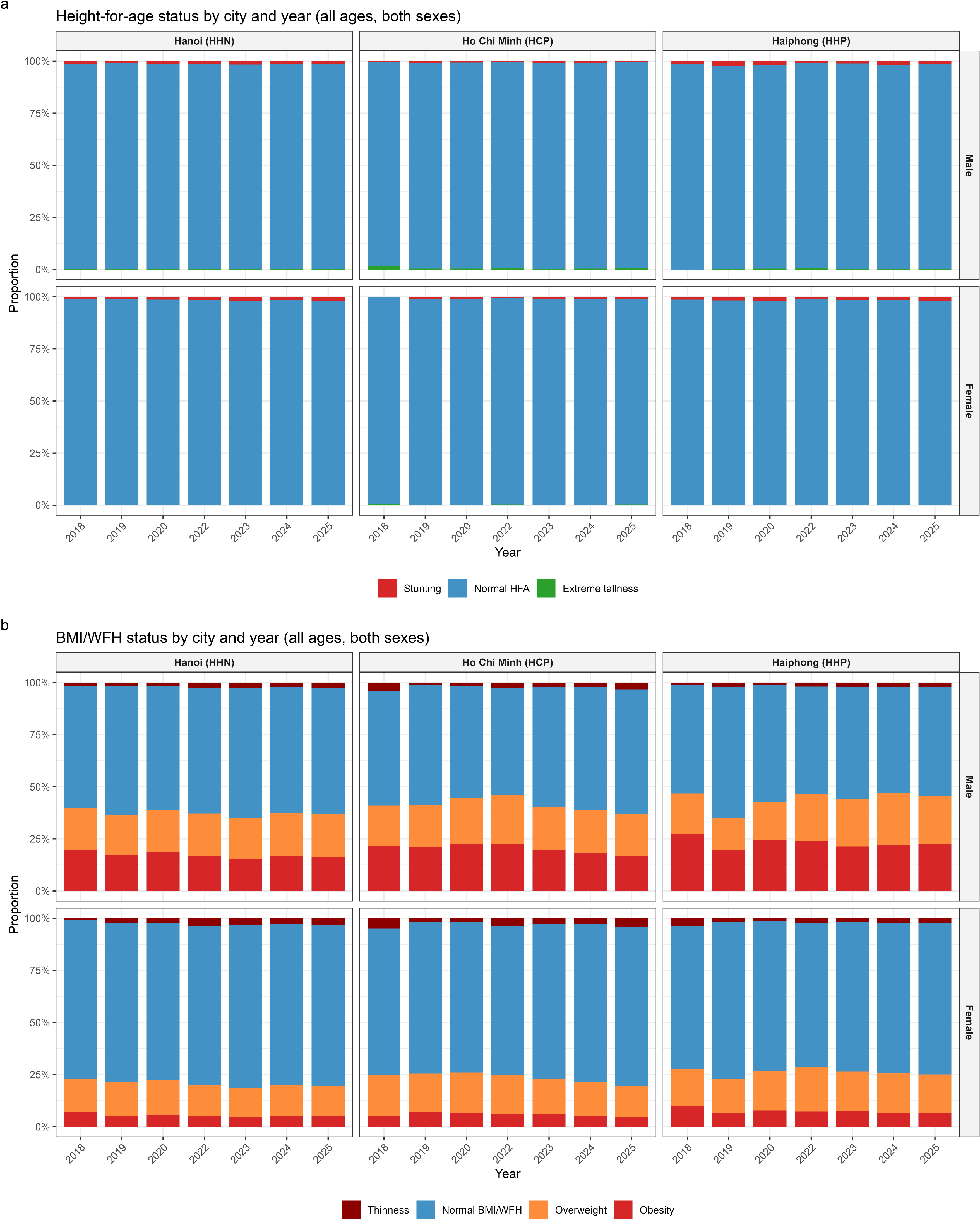
Stacked plot of nutritional status classification by city and sex. (a) Stacked proportional bar charts of height-for-age (HFA) nutritional status categories by city, calendar year, and sex. (b) Stacked proportional bar charts of BMI/WFH-for-age nutritional status categories by city, calendar year and sex. City labels: HHN = Hanoi, HCP = Ho Chi Minh City, HHP = Haiphong.

### Secular Trends in Thinness, Overweight, and Obesity

**Table 2** presents the annual prevalence of thinness, overweight, and obesity by sex. Thinness prevalence increased significantly in both sexes (males: *p* < 0.0001, females: *p* < 0.0001), rising from 3.0% (95%CI = 1.3, 4.7%) in males and 3.4% (95%CI = 1.9, 4.9%) in females in 2018 to 2.7% (95%CI = 2.1, 3.4%) and 3.5% (95%CI = 3.0, 4.1%) in 2025, respectively. However, the trajectory was not monotonic in that thinness fell substantially from 2018 to 2019 before rising progressively from 2020 to 2025 (**Figure 1**, **Figure 2**).

Overweight prevalence increased significantly across the study period in both sexes (males: *p* = 0.010, females: *p* < 0.0001). In males, overweight rose from 19.7% (95%CI= 18.0, 21.4%) in 2018 to 20.5% (95%CI =19.9, 21.1%) in 2025. In females, overweight declined slightly from 18.0% (95%CI =16.6, 19.6%) in 2018 to 14.8% (95%CI= 14.3, 15.4%) in 2025, a significant downward trend (*p* < 0.0001). Obesity showed highly significant trends in both sexes (both *p* < 0.0001), but in opposite directions. In males, obesity declined from 21.4% (95%CI =19.7, 23.1%) in 2018 to 17.0% (95%CI = 16.4, 17.6%) in 2025. In females, obesity also declined from 6.2% (95%CI = 4.8, 7.8%) in 2018 to 5.0% (95%CI = 4.5, 5.5%) in 2025 (**Figure** 1, Figure 3).

The GEE models confirmed divergent patterns by age group (**Figure 1**, **Figure 2**, **Figure 3**, **Table 2**). Thinness showed a significant annual increase in boys aged 5 to <10 years (OR= 1.133, 95%CI= 1.100, 1.167) and a significant annual decrease in boys aged 15 to 18 years (OR= 0.865, 95%CI= 0.807, 0.928). In girls aged 5 to <10 years, thinness also increased significantly (OR= 1.082, 95%CI= 1.053, 1.112). Obesity showed consistent significant annual decreases in the under 5 and 5 to <10-year groups for both sexes (male under 5: OR= 0.904, 95%CI= 0.869, 0.941, male 5 to <10 years: OR= 0.950, 95%CI= 0.939, 0.961, female under 5: OR= 0.906, 95%CI= 0.844, 0.972, female 5 to <10 years: OR= 0.964, 95%CI= 0.947, 0.981), but significant annual increases were observed among males aged 15 to 18 years (OR= 1.055, 95%CI= 1.006, 1.107). These age-group divergences suggest that while early-childhood obesity is declining, adolescent male obesity is rising.

### Double Burden of Malnutrition

**Figure 4** presents temporal trends in the double burden of malnutrition (DBM), and the full eight-category cross-tabulation is provided in **Supplementary Materials**. The predominant nutritional state throughout the study period was normal HFA combined with normal adiposity, which accounted for 54.9% of male and 71.4% of female observations in 2018. The category of normal HFA combined with overweight or obesity represented the next largest group, reflecting the predominantly non-stunting overweight phenotype characteristic of urban Vietnamese schoolchildren.

**Figure 4.**
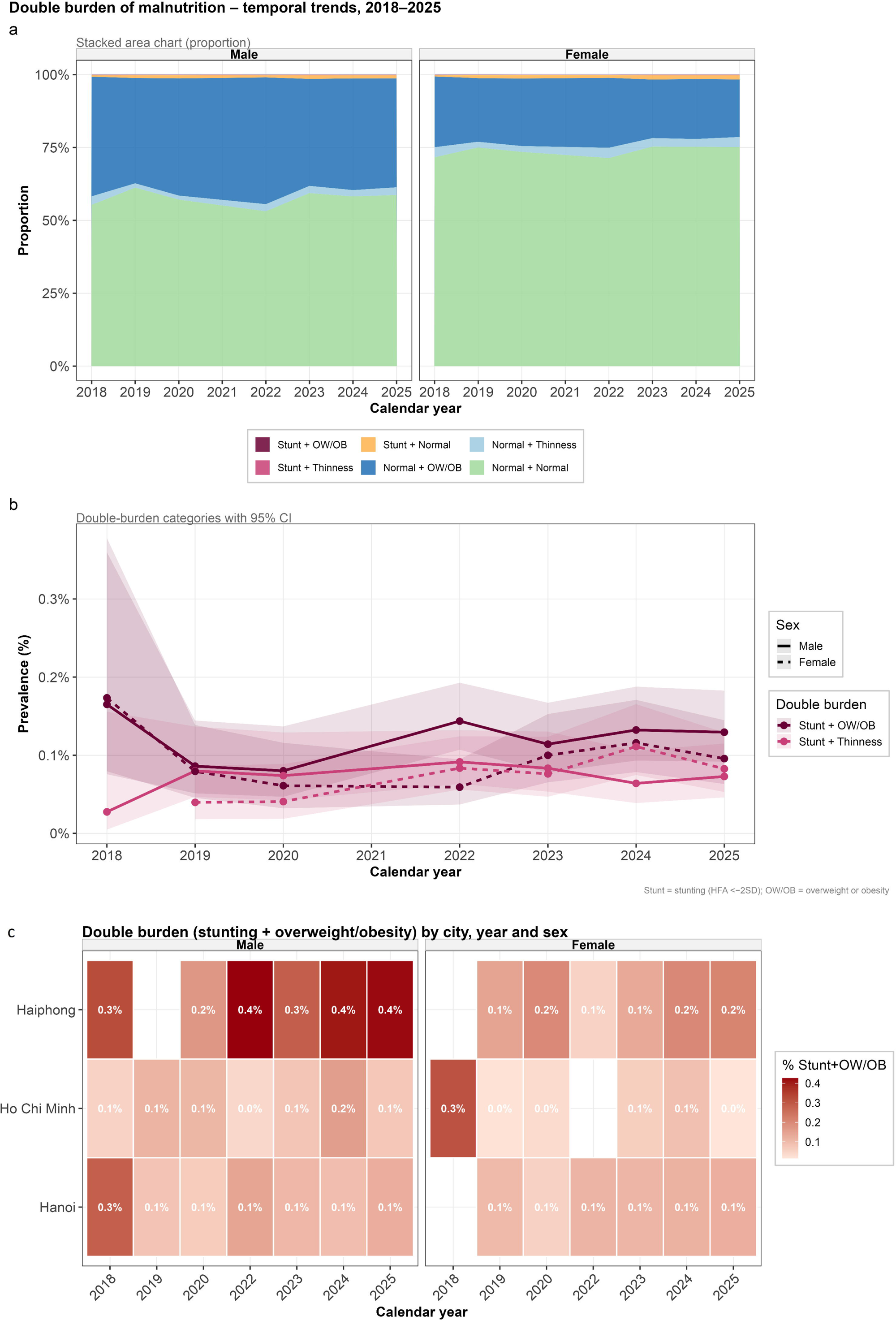
(a) Stacked area chart of double burden of malnutrition proportions by year and sex, 2018–2025. (b) Trend lines for the two clinically relevant double-burden categories (Stunting + OW/OB, Stunting + Thinness) with 95% CIs. OW/OB = overweight or obesity. (c) Heatmap of Stunting + Overweight/Obesity double-burden prevalence (%) by city, calendar year, and sex. Cell values represent annual proportions, color intensity indicates prevalence magnitude. City labels: HHN = Hanoi, HCP = Ho Chi Minh City, HHP = Haiphong.

The individual-level double burden of concurrent stunting with overweight or obesity (Stunting + OW/OB) was uncommon but detectable across years from 0.1% to 0.4%. The highest prevalence was observed in Haiphong (0.2% to 0.4% in males and 0.1% to 0.2% in females) and lowest in Hochiminh (around 0% to 0.2%) (**Figure 4**). The concurrent Stunting and Thinness category similarly remained rare (<0.1% throughout), consistent with the predominantly normal-to-overweight nutritional status of this school-based population.

### Regional Heterogeneity across Cities

Marked and persistent differences in nutritional status patterns were observed across the three cities. Ho Chi Minh City consistently showed the lowest stunting prevalence throughout (0.3 to 1.3% in males across all years), while Haiphong had the highest (1.0 to 2.2%). Haiphong showed the highest male obesity prevalence in 2025 (22.7%), compared with Hanoi (16.5%) and Ho Chi Minh City (16.8%) and also the highest prevalence of DBM of stunting and obesity (**Figures 3**, **Figure 4, Supplementary Materials**).

## DISCUSSION

### Principal Findings

In this 2018-2025 longitudinal annual school health check data of children aged 18 months to 18 years attending a private school system from 3 major cities in Vietnam, we document a complex and divergent nutritional transition characterized by three simultaneous trends including: a significant increase in stunting in the youngest age groups, an overall decline in early-childhood obesity, accompanied by a paradoxical significant increase in obesity among adolescent males. The individual-level double burden of concurrent stunting and overweight or obesity remained rare in this predominantly urban school-based population. Substantial and persistent city-level heterogeneity was observed across all outcomes, with Ho Chi Minh City showing lowest stunting prevalence, and Haiphong displaying highest stunting and obesity double burden. Taken together, these findings reveal an age-specific, and geographically heterogeneous nutritional epidemiology requiring targeted policy responses.

### Stunting in the Context of Vietnam’s Nutritional Progress

The prevalence of stunting in our data is remarkably lower than other reported data in Vietnam. Our observation of significant increases in stunting prevalence in children aged under 5 and 5 to <10 years (from 0.7% in both sexes in 2018 to 1.3% in males and 1.7% in females by 2025) appears counter-intuitive against Vietnam’s well-documented national narrative of stunting decline, where under-five stunting fell from 41.5% in 2000 to 19.3% by 2020 (12). Several explanations merit consideration. Our data is from a school-based cohort representing predominantly urban, economically advantaged households. The observed rising trend may reflect diet-induced stunting independent of caloric deficit, a phenomenon increasingly documented among children consuming energy-dense but micronutrient-poor diets in middle-income country urban settings (2,23).

The age-group specificity of the rising stunting trend which is concentrated in under 5 and 5 to <10-year groups with GEE odds ratios of 1.076 to 1.089 per additional calendar year, but non-significant in older adolescents is consistent with the critical-window hypothesis of growth faltering, which posits that stunting is predominantly determined by nutritional exposures in the first 1000 days and early childhood, after which the height trajectory is relatively fixed (24). The female-specific steeper increase (143% relative rise versus 86% in males), and the female excess in stunting emerging from 2023 onwards, is noteworthy and has not been previously reported in the Vietnamese urban surveillance literature. A Young Lives cohort analysis tracking longitudinal stunting transitions from childhood to adolescence confirmed that socioeconomic deterioration disproportionately drives stunting reversion in females in LMIC settings, providing a plausible mechanism for the observed sex divergence in our data (25).

Regarding thinness, a recent nationally representative cross-sectional survey of Vietnamese school-age children reported a national thinness prevalence of 5.1% (26), considerably higher than our 2025 estimate of 2.7 to 3.5%, consistent with the expectation that our data, drawn from predominantly middle-to-upper-income urban families, would have lower thinness than nationally representative samples inclusive of rural and lower-income populations.

### Divergent Obesity Trends by Age

The overall decline in obesity in males (from 21.4% to 17.0%, a relative reduction of 20.6%) and females (from 6.2% to 5.0%, a 19.4% decline) over 2018 to 2025 represents a noteworthy public health achievement, particularly given the pronounced rises in childhood obesity documented across middle-income countries over the same period (1,27). The decline was concentrated in the under 5 and 5 to <10-year groups, and may reflect a cohort effect whereby successive birth cohorts in urban Vietnam are experiencing lower early-onset obesity, potentially attributable to increased parental awareness, changes in infant feeding practices, or the effects of school meal improvement.

Against the overall declining trend, the significant annual increase in obesity among adolescent males (15 to 18 years: OR 1.055, p = 0.021) is a concerning signal. Adolescent obesity is more resistant to intervention than early-childhood obesity and more strongly associated with adult cardiometabolic risk (5). The absence of a corresponding significant increase in adolescent female obesity suggests sex-specific drivers, possibly including greater male engagement with energy-dense street foods, lower rates of dietary restraint, and differential responses to screen-time-related sedentary behavior. The consistently higher male obesity prevalence across all years and cities is consistent with the well-documented male excess in childhood obesity across Asia, attributed to sex-specific differences in body composition, dietary behavior, and physical activity (28).

### The Individual-Level Double Burden

The individual-level double burden of concurrent stunting with overweight or obesity (Stunting + OW/OB) was documented at 0.09 to 0.17% throughout the study period, without significant secular trend. Household-level DBM analyses across eight South and Southeast Asian countries documented prevalences of 10–12% (10), and trends analyses confirm that this form of double burden is rising in several ASEAN countries despite declining stunting at the national level (9), underscoring that economic development does not automatically resolve the coexistence of nutritional deficiency and excess. While low in absolute terms, our findings must be interpreted in context: our data represents predominantly advantaged urban schoolchildren in whom individual-level DBM is expected to be far less prevalent than in nationally representative samples or peri-urban populations.

### Strengths and Limitations

The notable strength of this study is the largest school-based longitudinal anthropometric dataset in Vietnam spanning eight calendar years across 3 cities, enabling characterization of secular trends and between city heterogeneity with substantially greater statistical power than previously available cross-sectional surveys in Vietnam (26).

There are some important limitations. First, the data were from private school students representing urban, predominantly economically advantaged families. As such the findings cannot be generalized to rural populations, ethnic minorities, or economically marginalized children who bear the heaviest stunting and thinness burden nationally (26). Second, the growing sample in later years as the private school network expanded may introduce differential selection effects. Third, the absence of dietary, physical activity, socioeconomic, or clinical data prevents causal analysis of the drivers of the observed trends. Fourth, the WHO growth references may not optimally represent Vietnamese healthy growth potential. However, this classification bias would be expected to remain stable over time and cannot explain the observed rising trends.

In summary, this study provides comprehensive longitudinal characterization of child nutritional status trends in urban Vietnam. The findings reveal a highly heterogeneous nutritional transition in which stunting and thinness are rising in younger children simultaneously with declining early-childhood obesity, a worsening adolescent male obesity burden, and marked city-level heterogeneity. The individual-level double burden of stunting and obesity are rare in this urban school-based population. These findings call for simultaneously targeted, age and sex specific and city-specific nutrition and school health policies to address various dimensions of malnutrition and overnutrition.

## Declarations

### Ethical Statement

The study was approved by the Vinmec Ethics Committee (approval number 0231/2024/CN/HDDD VMEC). Written informed consent was waived for secondary analysis of de-identified, retrospective data.

### Contributors

N.T.H. did conceptualization, data curation, formal analysis, investigation, methodology, project administration, resources, software, supervision, validation, visualization, writing original draft, and writing review & editing.

M.H., Q.N.N., L.N.P., and C.T.D. did supervision, writing review & editing.

M.A.N, Q.V.N, A.Q.D., C.T.L.T, H.T.M.L., T.V.P., L.T.P., and H.T.N. did data curation, resources, validation.

A.N.P did data curation, resources, supervision, validation, and writing– review & editing.

All authors read and approved the manuscript.

### Data Sharing Statement

Aggregated summary statistics and R code are available from the corresponding author upon reasonable request, subject to institutional ethics approval. Individual patient-level data cannot be shared due to applicable privacy regulations and the terms of the institutional ethics approval. The following will be made available upon reasonable request: anonymized aggregate data and analysis scripts.

### Use of Artificial Intelligence

All scientific content, analyses, and interpretations are the original work of the authors. The authors used AI-assisted tools for language editing and grammar checking during manuscript preparation.

## Abbreviations

BMI: body mass index
CI: confidence interval
DBM: double burden of malnutrition
GEE: generalized estimating equation
HFA: height-for-age
HCP: Ho Chi Minh City
HHN: Hanoi
HHP: Haiphong
LMIC: low- and middle-income country
OR: odds ratio
OW/OB: overweight or obesity
SD: standard deviation
WFH: weight-for-height
WHO: World Health Organization

