## Supplementary Materials for "Stunting, Thinness, Obesity, and Double Burden of Malnutrition in Vietnamese Urban Children"

Nhan Thi Ho et al.

**List of supplementary tables**

Table S1. Prevalence of stunting and tallness by year and sex.

Table S2. Prevalence of thinness, overweight, obesity by year and sex.

Table S3. Full prevalence of nutritional classification by year, city, sex.

Table S4. Temporal trend analysis form Generalized Estimation Equation (GEE) models.

Table S5. Cochrane-Armitage test for temporal trends.

Table S6. Prevalence of double burden categories by sex and year.

Table S1. Prevalence of stunting and tallness by year and sex.

| **Year visit** | **Sex** | **N total** | **N stunting** | **Percent stunting (95%CI)** | **N tallness** | **Percent tallness (95%CI)** |
| --- | --- | --- | --- | --- | --- | --- |
| 2018 | Male | 3546 | 26 | 0.7 (0.4, 1.1) | 34 | 1.0 (0.6, 1.4) |
| 2018 | Female | 3368 | 22 | 0.7 (0.4, 1.0) | 14 | 0.4 (0.1, 0.7) |
| 2019 | Male | 14675 | 168 | 1.1 (1.0, 1.3) | 50 | 0.3 (0.2, 0.5) |
| 2019 | Female | 13431 | 162 | 1.2 (1.0, 1.4) | 15 | 0.1 (0.0, 0.3) |
| 2020 | Male | 16275 | 207 | 1.3 (1.1, 1.5) | 64 | 0.4 (0.2, 0.6) |
| 2020 | Female | 14749 | 194 | 1.3 (1.1, 1.5) | 34 | 0.2 (0.0, 0.4) |
| 2022 | Male | 24126 | 263 | 1.1 (0.9, 1.2) | 100 | 0.4 (0.3, 0.6) |
| 2022 | Female | 22269 | 272 | 1.2 (1.1, 1.4) | 60 | 0.3 (0.1, 0.4) |
| 2023 | Male | 22803 | 334 | 1.5 (1.5, 1.5) | 71 | 0.3 (0.3, 0.3) |
| 2023 | Female | 21035 | 354 | 1.7 (1.5, 1.9) | 41 | 0.2 (0.0, 0.4) |
| 2024 | Male | 23445 | 306 | 1.3 (1.2, 1.5) | 84 | 0.4 (0.2, 0.5) |
| 2024 | Female | 21606 | 328 | 1.5 (1.4, 1.7) | 37 | 0.2 (0.0, 0.3) |
| 2025 | Male | 24670 | 320 | 1.3 (1.2, 1.4) | 92 | 0.4 (0.2, 0.5) |
| 2025 | Female | 22943 | 380 | 1.7 (1.5, 1.8) | 48 | 0.2 (0.0, 0.4) |

Table S2. Prevalence of thinness, overweight, obesity by year and sex.

| **Year visit** | **Sex** | **N total** | **N thinness** | **Percent thinness (95%CI)** | **N over weight** | **Percent overweight (95%CI)** | **N Obesity** | **Percent Obesity (95%CI)** |
| --- | --- | --- | --- | --- | --- | --- | --- | --- |
| 2018 | Male | 3542 | 106 | 3.0 (1.3, 4.7) | 698 | 19.7 (18.0, 21.4) | 758 | 21.4 (19.7, 23.1) |
| 2018 | Female | 3365 | 115 | 3.4 (1.9, 4.9) | 607 | 18.0 (16.6, 19.6) | 210 | 6.2 (4.8, 7.8) |
| 2019 | Male | 14664 | 235 | 1.6 (0.8, 2.4) | 2794 | 19.1 (18.2, 19.9) | 2674 | 18.2 (17.4, 19.1) |
| 2019 | Female | 13424 | 271 | 2.0 (1.3, 2.7) | 2259 | 16.8 (16.1, 17.6) | 767 | 5.7 (5.0, 6.4) |
| 2020 | Male | 16197 | 238 | 1.5 (0.7, 2.3) | 3297 | 20.4 (19.6, 21.2) | 3221 | 19.9 (19.1, 20.7) |
| 2020 | Female | 14700 | 303 | 2.1 (1.4, 2.8) | 2534 | 17.2 (16.5, 17.9) | 881 | 6.0 (5.3, 6.7) |
| 2022 | Male | 23625 | 621 | 2.6 (2.0, 3.3) | 5020 | 21.2 (20.6, 21.9) | 4548 | 19.3 (18.6, 19.9) |
| 2022 | Female | 21861 | 818 | 3.7 (3.2, 4.3) | 3605 | 16.5 (15.9, 17.1) | 1252 | 5.7 (5.2, 6.3) |
| 2023 | Male | 22783 | 586 | 2.6 (1.9, 3.2) | 4570 | 20.1 (19.4, 20.7) | 3815 | 16.7 (16.1, 17.4) |
| 2023 | Female | 21012 | 630 | 3.0 (2.4, 3.6) | 3167 | 15.1 (14.5, 15.6) | 1071 | 5.1 (4.5, 5.7) |
| 2024 | Male | 23428 | 528 | 2.3 (1.6, 2.9) | 4873 | 20.8 (20.1, 21.5) | 4130 | 17.6 (17.0, 18.3) |
| 2024 | Female | 21588 | 598 | 2.8 (2.2, 3.3) | 3332 | 15.4 (14.9, 16.0) | 1129 | 5.2 (4.7, 5.8) |
| 2025 | Male | 24658 | 671 | 2.7 (2.1, 3.4) | 5053 | 20.5 (19.9, 21.1) | 4194 | 17.0 (16.4, 17.6) |
| 2025 | Female | 22928 | 809 | 3.5 (3.0, 4.1) | 3404 | 14.8 (14.3, 15.4) | 1144 | 5.0 (4.5, 5.5) |

Table S3. Full prevalence of nutritional classification by year, city, sex.

| **Year visit** | **Hospital code** | **sex** | **N total** | **N stunting** | **% stunting** | **N total y** | **N thinness** | **N over weight** | **N obesity** | **% thinness** | **% over weight** | **% obesity** |
| --- | --- | --- | --- | --- | --- | --- | --- | --- | --- | --- | --- | --- |
| 2018 | HHN | Male | 1399 | 17 | 1.2 | 1399 | 25 | 282 | 277 | 1.8 | 20.2 | 19.8 |
| 2018 | HHN | Female | 1217 | 12 | 1 | 1217 | 12 | 193 | 85 | 1 | 15.9 | 7 |
| 2018 | HCP | Male | 1836 | 5 | 0.3 | 1833 | 77 | 356 | 396 | 4.2 | 19.4 | 21.6 |
| 2018 | HCP | Female | 1857 | 6 | 0.3 | 1854 | 92 | 362 | 96 | 5 | 19.5 | 5.2 |
| 2018 | HHP | Male | 311 | 4 | 1.3 | 310 | 4 | 60 | 85 | 1.3 | 19.4 | 27.4 |
| 2018 | HHP | Female | 294 | 4 | 1.4 | 294 | 11 | 52 | 29 | 3.7 | 17.7 | 9.9 |
| 2019 | HHN | Male | 11089 | 122 | 1.1 | 11081 | 188 | 2105 | 1926 | 1.7 | 19 | 17.4 |
| 2019 | HHN | Female | 9800 | 123 | 1.3 | 9795 | 204 | 1602 | 512 | 2.1 | 16.4 | 5.2 |
| 2019 | HCP | Male | 2997 | 33 | 1.1 | 2994 | 35 | 597 | 633 | 1.2 | 19.9 | 21.1 |
| 2019 | HCP | Female | 3112 | 30 | 1 | 3110 | 57 | 570 | 222 | 1.8 | 18.3 | 7.1 |
| 2019 | HHP | Male | 589 | 13 | 2.2 | 589 | 12 | 92 | 115 | 2 | 15.6 | 19.5 |
| 2019 | HHP | Female | 519 | 9 | 1.7 | 519 | 10 | 87 | 33 | 1.9 | 16.8 | 6.4 |
| 2020 | HHN | Male | 12285 | 166 | 1.4 | 12267 | 180 | 2471 | 2317 | 1.5 | 20.1 | 18.9 |
| 2020 | HHN | Female | 10821 | 146 | 1.3 | 10799 | 235 | 1789 | 606 | 2.2 | 16.6 | 5.6 |
| 2020 | HCP | Male | 2824 | 18 | 0.6 | 2765 | 43 | 613 | 619 | 1.6 | 22.2 | 22.4 |
| 2020 | HCP | Female | 2926 | 27 | 0.9 | 2900 | 54 | 557 | 197 | 1.9 | 19.2 | 6.8 |
| 2020 | HHP | Male | 1166 | 23 | 2 | 1165 | 15 | 213 | 285 | 1.3 | 18.3 | 24.5 |
| 2020 | HHP | Female | 1002 | 21 | 2.1 | 1001 | 14 | 188 | 78 | 1.4 | 18.8 | 7.8 |
| 2022 | HHN | Male | 15108 | 216 | 1.4 | 14756 | 398 | 2982 | 2504 | 2.7 | 20.2 | 17 |
| 2022 | HHN | Female | 13514 | 200 | 1.5 | 13251 | 516 | 1931 | 695 | 3.9 | 14.6 | 5.2 |
| 2022 | HCP | Male | 6619 | 24 | 0.4 | 6476 | 177 | 1502 | 1472 | 2.7 | 23.2 | 22.7 |
| 2022 | HCP | Female | 6637 | 47 | 0.7 | 6493 | 254 | 1220 | 403 | 3.9 | 18.8 | 6.2 |
| 2022 | HHP | Male | 2399 | 23 | 1 | 2393 | 46 | 536 | 572 | 1.9 | 22.4 | 23.9 |
| 2022 | HHP | Female | 2118 | 25 | 1.2 | 2117 | 48 | 454 | 154 | 2.3 | 21.4 | 7.3 |
| 2023 | HHN | Male | 15983 | 274 | 1.7 | 15972 | 436 | 3128 | 2437 | 2.7 | 19.6 | 15.3 |
| 2023 | HHN | Female | 14510 | 276 | 1.9 | 14496 | 464 | 2035 | 660 | 3.2 | 14 | 4.6 |
| 2023 | HCP | Male | 5084 | 40 | 0.8 | 5077 | 114 | 1043 | 1008 | 2.2 | 20.5 | 19.9 |
| 2023 | HCP | Female | 5025 | 57 | 1.1 | 5018 | 138 | 847 | 299 | 2.8 | 16.9 | 6 |
| 2023 | HHP | Male | 1736 | 20 | 1.2 | 1734 | 36 | 399 | 370 | 2.1 | 23 | 21.3 |
| 2023 | HHP | Female | 1500 | 21 | 1.4 | 1498 | 28 | 285 | 112 | 1.9 | 19 | 7.5 |
| 2024 | HHN | Male | 15926 | 220 | 1.4 | 15914 | 364 | 3227 | 2700 | 2.3 | 20.3 | 17 |
| 2024 | HHN | Female | 14457 | 233 | 1.6 | 14445 | 397 | 2116 | 749 | 2.7 | 14.6 | 5.2 |
| 2024 | HCP | Male | 5789 | 56 | 1 | 5784 | 125 | 1217 | 1045 | 2.2 | 21 | 18.1 |
| 2024 | HCP | Female | 5656 | 71 | 1.3 | 5652 | 168 | 933 | 281 | 3 | 16.5 | 5 |
| 2024 | HHP | Male | 1730 | 30 | 1.7 | 1730 | 39 | 429 | 385 | 2.3 | 24.8 | 22.3 |
| 2024 | HHP | Female | 1493 | 24 | 1.6 | 1491 | 33 | 283 | 99 | 2.2 | 19 | 6.6 |
| 2025 | HHN | Male | 16296 | 257 | 1.6 | 16286 | 420 | 3319 | 2688 | 2.6 | 20.4 | 16.5 |
| 2025 | HHN | Female | 14927 | 291 | 1.9 | 14917 | 505 | 2164 | 747 | 3.4 | 14.5 | 5 |
| 2025 | HCP | Male | 6705 | 38 | 0.6 | 6703 | 218 | 1353 | 1127 | 3.3 | 20.2 | 16.8 |
| 2025 | HCP | Female | 6570 | 63 | 1 | 6566 | 270 | 976 | 299 | 4.1 | 14.9 | 4.6 |
| 2025 | HHP | Male | 1669 | 25 | 1.5 | 1669 | 33 | 381 | 379 | 2 | 22.8 | 22.7 |
| 2025 | HHP | Female | 1446 | 26 | 1.8 | 1445 | 34 | 264 | 98 | 2.4 | 18.3 | 6.8 |

Table S4. Temporal trend analysis form Generalized Estimation Equation (GEE) models.

| **Age Category** | **Model** | **Annual trend Odds ratio (95%CI)** |
| --- | --- | --- |
| <5y | Stunting - Male | 1.076 (1.039, 1.116) |
| 5 to <10y | Stunting - Male | 1.053 (1.002, 1.108) |
| 10 to <15y | Stunting - Male | 1.029 (0.938, 1.129) |
| 15 to 18y | Stunting - Male | 0.973 (0.859, 1.102) |
| <5y | Stunting - Female | 1.089 (1.047, 1.134) |
| 5 to <10y | Stunting - Female | 1.086 (1.025, 1.150) |
| 10 to <15y | Stunting - Female | 1.022 (0.959, 1.089) |
| 15 to 18y | Stunting - Female | 1.098 (0.993, 1.213) |
| <5y | Thinness - Male | 1.050 (1.002, 1.100) |
| 5 to <10y | Thinness - Male | 1.133 (1.100, 1.167) |
| 10 to <15y | Thinness - Male | 1.026 (0.987, 1.067) |
| 15 to 18y | Thinness - Male | 0.865 (0.807, 0.928) |
| <5y | Thinness - Female | 0.982 (0.928, 1.040) |
| 5 to <10y | Thinness - Female | 1.082 (1.053, 1.112) |
| 10 to <15y | Thinness - Female | 1.044 (1.010, 1.079) |
| 15 to 18y | Thinness - Female | 1.028 (0.962, 1.099) |
| <5y | Obesity - Male | 0.904 (0.869, 0.941) |
| 5 to <10y | Obesity - Male | 0.950 (0.939, 0.961) |
| 10 to <15y | Obesity - Male | 0.994 (0.980, 1.008) |
| 15 to 18y | Obesity - Male | 1.055 (1.006, 1.107) |
| <5y | Obesity - Female | 0.906 (0.844, 0.972) |
| 5 to <10y | Obesity - Female | 0.964 (0.947, 0.981) |
| 10 to <15y | Obesity - Female | 0.997 (0.971, 1.023) |
| 15 to 18y | Obesity - Female | 1.079 (0.989, 1.178) |

Table S5. Cochrane-Armitage test for temporal trends.

| **Stratifier** | **Level** | **Outcome** | **Trend_p** |
| --- | --- | --- | --- |
| sex | Male | Stunting | 0.0063 |
| sex | Female | Stunting | 0 |
| sex | Male | Extreme tallness | 0.0265 |
| sex | Female | Extreme tallness | 0.541 |
| hospitalcode | HHN | Stunting | 0 |
| hospitalcode | HCP | Stunting | 0.0193 |
| hospitalcode | HHP | Stunting | 0.9608 |
| hospitalcode | HHN | Extreme tallness | 0.4956 |
| hospitalcode | HCP | Extreme tallness | 0.0179 |
| hospitalcode | HHP | Extreme tallness | 0.4081 |
| sex | Male | Thinness | 0 |
| sex | Female | Thinness | 0 |
| sex | Male | Overweight | 0.0098 |
| sex | Female | Overweight | 0 |
| sex | Male | Obesity | 0 |
| sex | Female | Obesity | 0 |
| hospitalcode | HHN | Thinness | 0 |
| hospitalcode | HCP | Thinness | 0.001 |
| hospitalcode | HHP | Thinness | 0.2978 |
| hospitalcode | HHN | Overweight | 0.0382 |
| hospitalcode | HCP | Overweight | 0 |
| hospitalcode | HHP | Overweight | 0.001 |
| hospitalcode | HHN | Obesity | 0 |
| hospitalcode | HCP | Obesity | 0 |
| hospitalcode | HHP | Obesity | 0.1426 |

Table S6. Prevalence of double burden categories by sex and year.

| **Year visit** | **Sex** | **Height for age (HFA) category** | **Body Mass Index (BMI) / Weight for height (WFH) category** | **N (%)** |
| --- | --- | --- | --- | --- |
| 2018 | Male | Extreme tallness | Normal BMI/WFH | 14 (0.39%) |
| 2018 | Male | Extreme tallness | Obesity | 10 (0.28%) |
| 2018 | Male | Extreme tallness | Overweight | 9 (0.25%) |
| 2018 | Male | Extreme tallness | Thinness | 1 (0.03%) |
| 2018 | Male | Normal HFA | Normal BMI/WFH | 1996 (54.93%) |
| 2018 | Male | Normal HFA | Obesity | 768 (21.13%) |
| 2018 | Male | Normal HFA | Overweight | 704 (19.37%) |
| 2018 | Male | Normal HFA | Thinness | 106 (2.92%) |
| 2018 | Male | Stunting | Normal BMI/WFH | 19 (0.52%) |
| 2018 | Male | Stunting | Obesity | 2 (0.06%) |
| 2018 | Male | Stunting | Overweight | 4 (0.11%) |
| 2018 | Male | Stunting | Thinness | 1 (0.03%) |
| 2018 | Female | Extreme tallness | Normal BMI/WFH | 8 (0.23%) |
| 2018 | Female | Extreme tallness | Obesity | 1 (0.03%) |
| 2018 | Female | Extreme tallness | Overweight | 2 (0.06%) |
| 2018 | Female | Extreme tallness | Thinness | 2 (0.06%) |
| 2018 | Female | Normal HFA | Normal BMI/WFH | 2471 (71.42%) |
| 2018 | Female | Normal HFA | Obesity | 212 (6.13%) |
| 2018 | Female | Normal HFA | Overweight | 624 (18.03%) |
| 2018 | Female | Normal HFA | Thinness | 118 (3.41%) |
| 2018 | Female | Stunting | Normal BMI/WFH | 16 (0.46%) |
| 2018 | Female | Stunting | Obesity | 3 (0.09%) |
| 2018 | Female | Stunting | Overweight | 3 (0.09%) |
| 2019 | Male | Extreme tallness | Normal BMI/WFH | 10 (0.06%) |
| 2019 | Male | Extreme tallness | Obesity | 23 (0.14%) |
| 2019 | Male | Extreme tallness | Overweight | 8 (0.05%) |
| 2019 | Male | Extreme tallness | Thinness | 5 (0.03%) |
| 2019 | Male | Normal HFA | Normal BMI/WFH | 9954 (61.17%) |
| 2019 | Male | Normal HFA | Obesity | 2852 (17.53%) |
| 2019 | Male | Normal HFA | Overweight | 2996 (18.41%) |
| 2019 | Male | Normal HFA | Thinness | 237 (1.46%) |
| 2019 | Male | Stunting | Normal BMI/WFH | 160 (0.98%) |
| 2019 | Male | Stunting | Obesity | 8 (0.05%) |
| 2019 | Male | Stunting | Overweight | 6 (0.04%) |
| 2019 | Male | Stunting | Thinness | 13 (0.08%) |
| 2019 | Female | Extreme tallness | Normal BMI/WFH | 5 (0.03%) |
| 2019 | Female | Extreme tallness | Obesity | 1 (0.01%) |
| 2019 | Female | Extreme tallness | Overweight | 3 (0.02%) |
| 2019 | Female | Extreme tallness | Thinness | 2 (0.01%) |
| 2019 | Female | Normal HFA | Normal BMI/WFH | 11335 (74.97%) |
| 2019 | Female | Normal HFA | Obesity | 858 (5.67%) |
| 2019 | Female | Normal HFA | Overweight | 2435 (16.1%) |
| 2019 | Female | Normal HFA | Thinness | 295 (1.95%) |
| 2019 | Female | Stunting | Normal BMI/WFH | 168 (1.11%) |
| 2019 | Female | Stunting | Obesity | 5 (0.03%) |
| 2019 | Female | Stunting | Overweight | 7 (0.05%) |
| 2019 | Female | Stunting | Thinness | 6 (0.04%) |
| 2020 | Male | Extreme tallness | Normal BMI/WFH | 10 (0.06%) |
| 2020 | Male | Extreme tallness | Obesity | 33 (0.2%) |
| 2020 | Male | Extreme tallness | Overweight | 12 (0.07%) |
| 2020 | Male | Extreme tallness | Thinness | 1 (0.01%) |
| 2020 | Male | Normal HFA | Normal BMI/WFH | 9267 (57.07%) |
| 2020 | Male | Normal HFA | Obesity | 3197 (19.69%) |
| 2020 | Male | Normal HFA | Overweight | 3286 (20.24%) |
| 2020 | Male | Normal HFA | Thinness | 225 (1.39%) |
| 2020 | Male | Stunting | Normal BMI/WFH | 183 (1.13%) |
| 2020 | Male | Stunting | Obesity | 4 (0.02%) |
| 2020 | Male | Stunting | Overweight | 9 (0.06%) |
| 2020 | Male | Stunting | Thinness | 12 (0.07%) |
| 2020 | Female | Extreme tallness | Normal BMI/WFH | 10 (0.07%) |
| 2020 | Female | Extreme tallness | Obesity | 11 (0.07%) |
| 2020 | Female | Extreme tallness | Overweight | 7 (0.05%) |
| 2020 | Female | Normal HFA | Normal BMI/WFH | 10831 (73.42%) |
| 2020 | Female | Normal HFA | Obesity | 870 (5.9%) |
| 2020 | Female | Normal HFA | Overweight | 2528 (17.14%) |
| 2020 | Female | Normal HFA | Thinness | 299 (2.03%) |
| 2020 | Female | Stunting | Normal BMI/WFH | 181 (1.23%) |
| 2020 | Female | Stunting | Obesity | 4 (0.03%) |
| 2020 | Female | Stunting | Overweight | 5 (0.03%) |
| 2020 | Female | Stunting | Thinness | 6 (0.04%) |
| 2022 | Male | Extreme tallness | Normal BMI/WFH | 26 (0.08%) |
| 2022 | Male | Extreme tallness | Obesity | 54 (0.18%) |
| 2022 | Male | Extreme tallness | Overweight | 35 (0.11%) |
| 2022 | Male | Extreme tallness | Thinness | 1 (0%) |
| 2022 | Male | Normal HFA | Normal BMI/WFH | 16241 (53.04%) |
| 2022 | Male | Normal HFA | Obesity | 6405 (20.92%) |
| 2022 | Male | Normal HFA | Overweight | 6820 (22.27%) |
| 2022 | Male | Normal HFA | Thinness | 749 (2.45%) |
| 2022 | Male | Stunting | Normal BMI/WFH | 220 (0.72%) |
| 2022 | Male | Stunting | Obesity | 22 (0.07%) |
| 2022 | Male | Stunting | Overweight | 22 (0.07%) |
| 2022 | Male | Stunting | Thinness | 28 (0.09%) |
| 2022 | Female | Extreme tallness | Normal BMI/WFH | 27 (0.09%) |
| 2022 | Female | Extreme tallness | Obesity | 17 (0.06%) |
| 2022 | Female | Extreme tallness | Overweight | 27 (0.09%) |
| 2022 | Female | Extreme tallness | Thinness | 2 (0.01%) |
| 2022 | Female | Normal HFA | Normal BMI/WFH | 20493 (71.29%) |
| 2022 | Female | Normal HFA | Obesity | 1724 (6%) |
| 2022 | Female | Normal HFA | Overweight | 5125 (17.83%) |
| 2022 | Female | Normal HFA | Thinness | 1016 (3.53%) |
| 2022 | Female | Stunting | Normal BMI/WFH | 272 (0.95%) |
| 2022 | Female | Stunting | Obesity | 2 (0.01%) |
| 2022 | Female | Stunting | Overweight | 15 (0.05%) |
| 2022 | Female | Stunting | Thinness | 24 (0.08%) |
| 2023 | Male | Extreme tallness | Normal BMI/WFH | 15 (0.07%) |
| 2023 | Male | Extreme tallness | Obesity | 35 (0.15%) |
| 2023 | Male | Extreme tallness | Overweight | 14 (0.06%) |
| 2023 | Male | Extreme tallness | Thinness | 4 (0.02%) |
| 2023 | Male | Normal HFA | Normal BMI/WFH | 13510 (59.3%) |
| 2023 | Male | Normal HFA | Obesity | 3771 (16.55%) |
| 2023 | Male | Normal HFA | Overweight | 4539 (19.92%) |
| 2023 | Male | Normal HFA | Thinness | 561 (2.46%) |
| 2023 | Male | Stunting | Normal BMI/WFH | 289 (1.27%) |
| 2023 | Male | Stunting | Obesity | 9 (0.04%) |
| 2023 | Male | Stunting | Overweight | 17 (0.07%) |
| 2023 | Male | Stunting | Thinness | 19 (0.08%) |
| 2023 | Female | Extreme tallness | Normal BMI/WFH | 18 (0.09%) |
| 2023 | Female | Extreme tallness | Obesity | 11 (0.05%) |
| 2023 | Female | Extreme tallness | Overweight | 7 (0.03%) |
| 2023 | Female | Extreme tallness | Thinness | 1 (0%) |
| 2023 | Female | Normal HFA | Normal BMI/WFH | 15811 (75.25%) |
| 2023 | Female | Normal HFA | Obesity | 1055 (5.02%) |
| 2023 | Female | Normal HFA | Overweight | 3144 (14.96%) |
| 2023 | Female | Normal HFA | Thinness | 611 (2.91%) |
| 2023 | Female | Stunting | Normal BMI/WFH | 317 (1.51%) |
| 2023 | Female | Stunting | Obesity | 5 (0.02%) |
| 2023 | Female | Stunting | Overweight | 16 (0.08%) |
| 2023 | Female | Stunting | Thinness | 16 (0.08%) |
| 2024 | Male | Extreme tallness | Normal BMI/WFH | 18 (0.08%) |
| 2024 | Male | Extreme tallness | Obesity | 42 (0.18%) |
| 2024 | Male | Extreme tallness | Overweight | 17 (0.07%) |
| 2024 | Male | Extreme tallness | Thinness | 4 (0.02%) |
| 2024 | Male | Normal HFA | Normal BMI/WFH | 13620 (58.14%) |
| 2024 | Male | Normal HFA | Obesity | 4078 (17.41%) |
| 2024 | Male | Normal HFA | Overweight | 4835 (20.64%) |
| 2024 | Male | Normal HFA | Thinness | 509 (2.17%) |
| 2024 | Male | Stunting | Normal BMI/WFH | 259 (1.11%) |
| 2024 | Male | Stunting | Obesity | 10 (0.04%) |
| 2024 | Male | Stunting | Overweight | 21 (0.09%) |
| 2024 | Male | Stunting | Thinness | 15 (0.06%) |
| 2024 | Female | Extreme tallness | Normal BMI/WFH | 13 (0.06%) |
| 2024 | Female | Extreme tallness | Obesity | 11 (0.05%) |
| 2024 | Female | Extreme tallness | Overweight | 8 (0.04%) |
| 2024 | Female | Extreme tallness | Thinness | 5 (0.02%) |
| 2024 | Female | Normal HFA | Normal BMI/WFH | 16239 (75.22%) |
| 2024 | Female | Normal HFA | Obesity | 1115 (5.16%) |
| 2024 | Female | Normal HFA | Overweight | 3302 (15.3%) |
| 2024 | Female | Normal HFA | Thinness | 569 (2.64%) |
| 2024 | Female | Stunting | Normal BMI/WFH | 277 (1.28%) |
| 2024 | Female | Stunting | Obesity | 3 (0.01%) |
| 2024 | Female | Stunting | Overweight | 22 (0.1%) |
| 2024 | Female | Stunting | Thinness | 24 (0.11%) |
| 2025 | Male | Extreme tallness | Normal BMI/WFH | 28 (0.11%) |
| 2025 | Male | Extreme tallness | Obesity | 41 (0.17%) |
| 2025 | Male | Extreme tallness | Overweight | 20 (0.08%) |
| 2025 | Male | Extreme tallness | Thinness | 1 (0%) |
| 2025 | Male | Normal HFA | Normal BMI/WFH | 14481 (58.6%) |
| 2025 | Male | Normal HFA | Obesity | 4147 (16.78%) |
| 2025 | Male | Normal HFA | Overweight | 5023 (20.33%) |
| 2025 | Male | Normal HFA | Thinness | 652 (2.64%) |
| 2025 | Male | Stunting | Normal BMI/WFH | 269 (1.09%) |
| 2025 | Male | Stunting | Obesity | 7 (0.03%) |
| 2025 | Male | Stunting | Overweight | 25 (0.1%) |
| 2025 | Male | Stunting | Thinness | 18 (0.07%) |
| 2025 | Female | Extreme tallness | Normal BMI/WFH | 19 (0.08%) |
| 2025 | Female | Extreme tallness | Obesity | 11 (0.05%) |
| 2025 | Female | Extreme tallness | Overweight | 14 (0.06%) |
| 2025 | Female | Extreme tallness | Thinness | 2 (0.01%) |
| 2025 | Female | Normal HFA | Normal BMI/WFH | 17260 (75.07%) |
| 2025 | Female | Normal HFA | Obesity | 1132 (4.92%) |
| 2025 | Female | Normal HFA | Overweight | 3378 (14.69%) |
| 2025 | Female | Normal HFA | Thinness | 796 (3.46%) |
| 2025 | Female | Stunting | Normal BMI/WFH | 339 (1.47%) |
| 2025 | Female | Stunting | Obesity | 3 (0.01%) |
| 2025 | Female | Stunting | Overweight | 19 (0.08%) |
| 2025 | Female | Stunting | Thinness | 19 (0.08%) |
